# Verification of the Abbott Alinity m Resp-4-Plex Assay for detection of SARS-CoV-2, influenza A/B, and respiratory syncytial virus

**DOI:** 10.1101/2021.04.22.21255133

**Authors:** Annie Cheng, Stefan Riedel, Ramy Arnaout, James E. Kirby

## Abstract

COVID-19 symptomology may overlap with other circulating respiratory viruses that may also cause severe disease and for which there are specific and potentially life-saving treatments. The Abbott Alinity m Resp-4-Plex assay is a multiplex PCR assay that simultaneously detects and differentiates infection with SARS-CoV-2, influenza A, influenza B, and respiratory syncytial virus (RSV). We characterized its accuracy, precision, and analytical sensitivity. All were found to be robust for measures examined. In the context of sample-to-answer, near random access automation on the Alinity m platform, we believe that the Resp-4-Plex assay offers significant utility in addressing the current needs of the SARS-CoV-2 pandemic and future needs during anticipated endemic circulation of SARS-CoV-2 with other respiratory viruses.

## Introduction

SARS-CoV-2, influenza A/B, and RSV may cause respiratory infection with significant morbidity and mortality. Respiratory disease signs and symptoms for these viruses overlap, and, therefore, it is not possible to reliably differentiate between them on clinical grounds alone, especially early during the course of disease. SARS-CoV-2 and influenza, especially, have significant implications in terms of transmission inside and outside of hospital settings and therefore require reliable methods for diagnosis. RSV, although primarily thought of as a serious pathogen in young children, can also cause bronchiolitis and pneumonia in adults. Depending on the stage of illness, there are therapeutics with varying efficacy for SARS-CoV-2, influenza, and RSV. Therefore, sensitive detection and differentiation of these viruses are valuable clinical determinations.

With social distancing and masking during the COVID pandemic, the circulation of influenza and other respiratory viruses almost ceased in many locations (1). Presumably, however, with less than 100% vaccine efficacy for influenza, waning immunity to circulating respiratory viruses over a large population cohort, and reopening of our societies, circulation of influenza and RSV will rebound and likely exceed normal levels for some time (2). At the same time, SARS-CoV-2 will likely become endemic, potentially adopting a seasonal cycle with enhanced transmission during the winter as observed in the United States during the winters of 2020 and 2021 (3). It will therefore be critical to be able to test both patients and staff for high consequence respiratory pathogens to avert potential nosocomial transmission and to identify the most advantageous therapeutic options for patients with serious illness.

A multiplex testing option for high consequence testing options would address these specific diagnostic needs. The Abbott Alinity m Resp-4-Plex assay in March 2021 received emergency use authorization designation for detection of SARS-CoV-2, influenza A, influenza B and RSV. Approved sample types are either a nasopharyngeal swab collected by a health care provider or a nasal swab specimen self-collected in a healthcare setting. The multiplex, reverse-transcription real-time PCR assay targets the RdRp and N genes of SARS-CoV2: the matrix gene of influenza A; the nonstructural 1 gene of influenza B, and the matrix gene of RSV. An internal control is spiked into each sample in the form of armored RNA encoding a segment of the hydroxypyruvate reductase gene from the pumpkin plant, *Cucurbita pepo*. It controls for appropriate extraction and amplification in each reaction. Each amplicon is detected by a real-time probe with a distinct fluorophore with the exception that both probes for the SARS-CoV-2 targets are detected with the same fluorophore. The primers and probes for the SARS-CoV-2 target are the same as those used in the singleplex Abbott RealTime SARS-CoV-2 and Alinity m SARS-CoV-2 assays, whose performance characteristics have been examined in prior literature (4-6). Cycle threshold numbers (Ct) determined on the Alinity m instrument in the Resp-4-Plex assay are determined based on a fluorescence cutoff. They are used along with the inflection point of the amplification curve at the maximum amplification efficiency (the max ratio) (7) for qualitative assessment of target positivity and negativity (personal communication, Joshua Kostera, Abbott Molecular).

Here we describe characterization of accuracy, precision, and limit of detection of the Alinity m Resp-4-Plex assay determined as part of normal quality assurance activities prior to adoption for clinical use in our clinical laboratory.

## Materials and Methods

### Accuracy

Results from prior determinations either by Cepheid Xpert® Xpress Flu; direct fluorescent antigen testing for influenza A/B, RSV, adenovirus, and parainfluenza 1, 2 and 3 and/or shell vial culture on R-Mix-Too monolayers (Quidel Corporation, San Diego, CA); and/or reference laboratory testing at Eurofin ViraCor using the TEM-PCR assay (8) were considered predicate comparator assays as enumerated. Samples were divided for Resp-4-Plex testing on the two Alinity m instruments at our institution. As there was no difference in performance on these identical systems results are presented in aggregate. Discrepant resolution included repeat testing of samples as available using Resp-4-Plex on the second Alinity m platform and/or using the Cepheid Xpert® Xpress SARS-CoV-2/Flu/RSV assay. Data for individual sample is listed in Table S1.

### Analytical sensitivity

For limit of detection studies, the AccuPlex SARS-CoV-2, Flu A/B and RSV Verification Panel (catalogue #0505-0183, LGC SeraCare, Milford, MA) with individual members at defined equal concentrations, quantified by digital droplet PCR, were initially diluted to 1E4, 5E3, 1E3, 5E2, 2E2, 1E2, 5E1, 2.5E1, 1E1 and 0.5E1 target amplicon genome copies per mL and tested by Resp-4-Plex in quadruplicate. Each panel member consists of either part or the entire genome of the target virus cloned into a replication incompetent Sindbis virus. The Sindbis virus is an enveloped, single-stranded, RNA genome virus that serves as a surrogate and control for all processes in the assay including extraction. The linearity of amplification of each assay was determined by least squares linear regression based on Ct values obtained from each screening concentration, and PCR efficiency was determined using slope of the regression line using the ThermoFisher calculator: https://www.thermofisher.com/us/en/home/brands/thermo-scientific/molecular-biology/molecular-biology-learning-center/molecular-biology-resource-library/thermo-scientific-web-tools/qpcr-efficiency-calculator.html. Three dilutions bracketing the potential limit of detection cutoff were tested again with twenty replicates each to establish the LoD for each analyte. The 95% detection threshold and confidence intervals were extrapolated by Logistic regression in Prism 9 for MacOS (GraphPad, San Diego, CA). LoD analysis was only performed on a single Alinity m instrument.

## Results

### Accuracy

To determine accuracy, the Resp-4-Plex assay was run using clinical samples that were previously determined to be positive for the constituent viruses. Ten samples previously determined to be SARS-CoV-2 positive using the Alinity m SARS-CoV-2 singleplex assay were tested using Resp-4-Plex. All were positive for SARS-CoV-2. Although the Ct values for SARS-CoV-2 on Resp-4-Plex were a mean −1.81 +/-0.58 lower than fractional cycle numbers on the singleplex assay, the assays were highly correlated with one another (R^2^ = 0.99) (Fig 1A).

**Figure 1.**
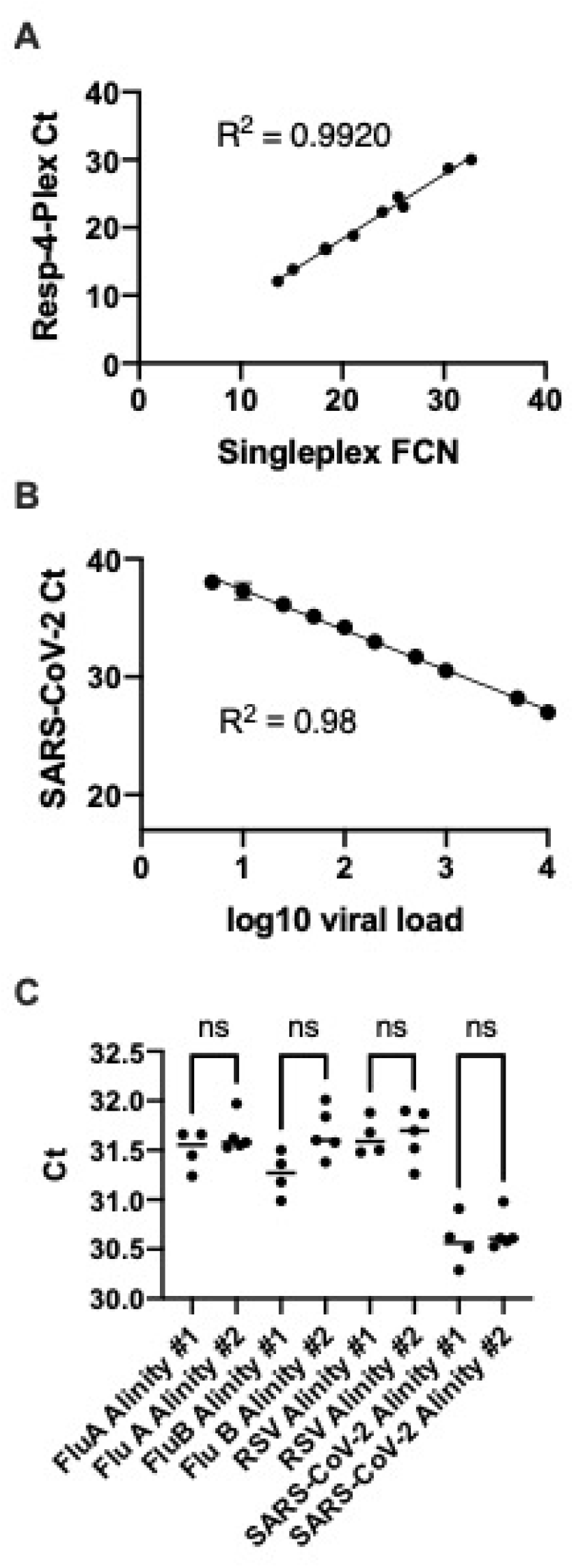
Performance of the Resp-4-Plex Assay. **(A)** Cycle threshold (Ct) results from the SARS-CoV-2 test in the Alinity m Resp-4-Plex multiplex assay and fractional cycle numbers (FCN) (7) from the Alinity m SARS-CoV-2 singleplex assay were highly correlated when testing patient samples spanning the analytical measurement range of these tests. **(B)** Amplification of the SARS-CoV-2 targets in the Resp-4-Plex assay was log-linear when examined in the range at and above the limit of detection. Data points shown are the mean and standard deviation of Ct values for 4 replicate measurements. **(C)** Data points for positive control Ct values for each individual assay in Resp-4-Plex, tested on four to five separate days on two separate Alinity instruments, were highly correlated and not statistically (n.s.) different between instruments.

Twelve samples previously determined to be influenza A positive by Cepheid Xpress Flu were tested using Resp-4-Plex. All tested positive influenza A. A single sample tested with Resp-4-Plex also yielded a positive RSV result with a high Ct value of 32.46 (see limit of detection analysis below). The test was repeated using Resp-4-Plex on the second Alinity m instrument and gave qualitatively and quantitatively almost identical results. The sample was then tested using the Cepheid Xpress SARS-CoV-2/Flu/RSV assay and was influenza A positive, RSV negative. This sample was interpreted as a potential Xpress RSV false negative based on low viral load.

Ten samples previously determined to be influenza B positive using the Cepheid Xpress Flu assay were tested using Resp-4-Plex. All tested positive for influenza B. A single sample tested with Resp-4-Plex also yield a positive RSV result. The test was repeated using Resp-4-Plex on the second Alinity m instrument and gave similar results. The sample was then tested using the Cepheid Xpress SARS-CoV-2/Flu/RSV assay and was influenza B positive, RSV positive. This sample was resolved as an influenza B/RSV true positive.

Ten samples previously determined to be RSV positive using a combination of direct fluorescence antigen testing and/or shell vial culture and a single sample determined to be RSV positive by TEM-PCR were tested using Resp-4-Plex. All were positive for RSV.

Eighteen samples previously determined to be negative for viruses by direct fluorescent antigen testing and/or shell vial culture, five of which were also negative by TEM-PCR, were tested using Resp-4-Plex. Sixteen samples were negative for the viruses detected by Resp-4-Plex. One sample tested with Resp-4-Plex yield as positive result for influenza B with a high Ct value of 34.2. The test was repeated using Resp-4-Plex on the second Alinity m instrument and gave similar results. The sample was then tested using the Cepheid Xpress SARS-CoV-2/Flu/RSV assay and was influenza B positive. A second sample (also negative by TEM-PCR) yielded a positive result for influenza A with Resp-4-Plex with a high Ct value of 36.53 and a positive result for RSV with a lower Ct value of 21.82. The test was repeated using Resp-4-Plex on the second Alinity m instrument and gave similar results. The sample was then tested using the Cepheid Xpress SARS-CoV-2/Flu/RSV assay and was influenza A negative (however, with a Ct value of 38.6) and RSV positive (Ct value of 23.4). The sample was resolved as likely true positive on Resp-4-Plex for influenza A and RSV.

Six samples positive for Adenovirus or Parainfluenza virus 1, 2, or 3 by direct fluorescent antigen testing, one sample positive for parainfluenza, two samples positive for adenovirus or parainfluenza virus by shell vial culture, 1 sample positive for enterovirus by TEM-PCR and three samples positive for low pathogenicity coronaviruses by TEM-PCR were tested using Resp-4-Plex. All samples were negative for the viruses detected by Resp-4-Plex.

### Analytical Sensitivity

For SARS-CoV-2, the LoD screen demonstrated excellent linearity with R^2^ = 0.98 and a 97% PCR efficiency (Fig. 1B). The manufacturer’s claimed LoD was confirmed through twenty replicates at 50, 25 and 10 copies/mL using the SeraCare reference material, yielding 100%, 100% and 80% detection, respectively. The LoD was therefore <= 25 copies per mL with an Ct value of 35.72 +/-0.54 at 25 copies/mL. The logistic regression was non-convergent and therefore an extrapolated LoD could not be established.

For influenza A, the LoD screen demonstrated excellent linearity with R^2^ = 0.97 and 93% PCR efficiency. The manufacturer’s claimed LoD was confirmed through twenty replicates at 50, 25 and 10 copies/mL using the SeraCare reference material, yielding 100%, 80% and 30% detection, respectively. The LoD was therefore <= to 50 copies per mL with an Ct value of 37.18+/-0.67 at 50 copies/mL. By logistic regression, the LoD was 47 copies/mL (95% confidence interval ∼25-125 copies/mL).

For influenza B, the LoD screen demonstrated excellent linearity with an R^2^ = 0.97 and a 101% PCR efficiency. The manufacturer’s claimed LoD was confirmed through twenty replicates at 50, 25 and 10 copies/mL using the SeraCare reference material, yielding 100%, 90% and 70% detection, respectively. The LoD was therefore <= to 50 copies per mL with an Ct value of 35.94+/-0.73 at 50 copies/mL. By logistic regression, the LoD was 36 copies/mL (95% confidence interval ∼18-178 copies/mL).

For RSV, the LoD screen demonstrated excellent linearity with an R^2^ = 0.97 and 97% PCR efficiency. The manufacturer’s claimed LoD was confirmed through twenty replicates at 50, 25 and 10 copies/mL using the SeraCare reference material, yielding 100%, 90% and 75% detection, respectively. The LoD was <= to 50 copies per mL with an Ct value of 35.75 +/-0.70 at 50 copies/mL. By logistic regression the LoD was 39.8 copies/mL (95% confidence interval ∼18-1000 copies/mL).

### Precision

Intra-run precision was determined by using pooled samples, positive for each of the four viruses, and a negative pool. Each pool was tested in quadruplicate on each of the two Alinity m instruments with each individual test for each specific virus performed on each of the Assay Processing Units (APU) #1 through #4 on each instrument, so that every APU was tested during the precision testing. Inter-run precision was verified by testing the pools for each virus on each alinity m instrument again on additional two separate days. Qualitative intra-run and inter-run precision results were 100% correlated as expected.

Although Resp-4-Plex is a qualitative assay, we also assessed quantitative precision by comparison of Ct values in replicates. Coefficients of variation (CV) varied from 0.6 to 2.5% across all viruses tested on both instruments in intra-run precision comparisons and from 0.4% to 2.9% in inter-run precision comparisons. The precision of the Ct values for positive controls run during the validation on Alinity m #1 (n=4) and Alinity m #2 (n=5) instruments, run once per day of testing, was also examined. C.V.’s on individual instruments for each assay were all less than 1%. There was no statistical difference between the positive control Ct values run on Alinity #1 and Alinity #2 for the four viruses (Fig. 1C), with significance considered *P* <= 0.05, with comparisons performed using the Kruskall-Wallis test. Therefore, reproducibility of both instruments appeared essentially identical for the Resp-4-Plex assay.

## Discussion

The Resp-4-Plex assay is welcome addition to targeted respiratory panel options that will be necessary in a post-COVID world. It appeared highly accurate, sensitive, and precise. The three descrepancies with comparator methods, specifically detection of co-infections, could be attributed to enhanced detection by Resp-4-Plex. This was because: (1) the second virus was detected on repeat testing by Resp-4-Plex on a second Alinity m platform, (2) the second virus was detected by the alternative Cepheid respiratory panel, and/or (3) the second virus had a very high Ct value, near its limit of detection, and therefore plausibly may have been below the limit of detection of the comparator assays. Overall, our experience was consistent with qualitative detection data described in the EUA product insert, although our data set was significantly smaller in size.

Notably, the LoD for each individual virus was robust with high amplification efficiencies even in the context of a multiplex assay and testing of quality control material in which all four targets were present in equivalent amounts. In fact, the limit of detection for SARS-CoV-2 in Resp-4-Plex (∼25 copies/mL) was below that determined in our analysis of the SARS-CoV-2 SinglePlex assay (∼50 copies/mL, data not shown). The multiplex and singleplex SARS-CoV-2 assays were also extremely well correlated (R^2^ = 0.99).

Furthermore, the Ct values were also log-linearly correlated with the quantitative standard at and above the limit of detection of the assays with excellent PCR efficiencies. Notably, Ct values or their equivalent (e.g. fractional cycle number (7)) may vary significantly between platforms at the same viral load level and for difference viruses (9). Furthermore, they vary inversely with viral load which is intuitively confusing to end users. We ultimately believe that SARS-CoV-2 results should be reported selectively as a viral load, as most intuitively understood by clinicians, benchmarked against a universal standard (6). In particular, there are several situations where viral load values are helpful, for example, to distinguish between reinfection versus persistent low levels of mRNA that may last months after a prior infection, and to evalute likely infectivity in the appropriate contexts. The log-linear performance of the Resp-4-Plex assays suggest that future conversion to a quantitative readout, i.e., a viral load, should easily be supported and we believe will provide additional utility for patient management. Furthermore, our previous study with the Abbott RealTime SARS-CoV-2 assay (6) and the singleplex Alinity m SARS-CoV-2 assay (data not shown) supports a method for accurately calculating the viral load from Ct measurements without the absolute need for a standard curve, and/or extending viral load determinations beyond limits of available calibrator material.

Taken together, we verified the performance characteristis of a new molecular multiplex respiratory panel assay on the Alinity m molecular system.. This platform notably provides high throughput; sample-to-answer, random access and semi-batch functionality with a 115 minute sample to answer turn around time for prioritized specimens; and an ability to load and perform multiple different tests at one time. We believe this assay and platform will be especially useful in fulfilling future needs, in situations where SARS-CoV-2, influenza A, influenza B and RSV circulate at significant levels, and where these viruses need to be detected and differentiated for optimal patient management.

## Data Availability

All data is available in manuscript and supplemental data.

## Acknowledgements

We thank the Abbott Molecular Group for providing test reagents and LGC SeraCare control material for this work under a COVID-19 Diagnostics Evaluation Agreement. RA was a recipient of grant support from Abbott Molecular under a clinical study agreement.

## Author Contribution Statement

Annie Cheng conceptualized and performed experiments. Stefan Riedel reviewed and edited manuscript and aided in formal analysis. Ramy Arnaout reviewed and edited the manuscript and aided in formal analysis. James Kirby wrote the initial manuscript draft and aided in conceptualization and formal analysis.

## Notes

### Author Declarations

The study was performed as part of normal quality assurance/quality improvement (QI) activities in our clinical laboratory. The BIDMC IRB reviewed the proposed activity and determined that the study does not constitute human subjects research as stated in its determination letter.

